# Insomnia and risk of sepsis: A Mendelian randomization study

**DOI:** 10.1101/2022.09.02.22279472

**Authors:** Marianne S. Thorkildsen, Lise T. Gustad, Randi M. Mohus, Tom I.L. Nilsen, Jan K. Damås, Tormod Rogne

**Affiliations:** Gemini Center for Sepsis Research at Institute of Circulation and Medical Imaging, NTNU, Trondheim, Norway; Faculty of Nursing and Health Sciences, Nord University, Levanger, Norway; Department of Medicine and Rehabilitation, Levanger Hospital, Nord-Trøndelag Hospital Trust, Levanger, Norway; Clinic of Anesthesia and Intensive Care, St. Olavs hospital, Trondheim, Norway; Department of Circulation and Medical Imaging, NTNU, Trondheim, Norway; Department of Public Health and Nursing, NTNU, Trondheim, Norway; Centre of Molecular Inflammation Research, NTNU, Trondheim, Norway; Department of Clinical and Molecular Medicine, NTNU, Trondheim, Norway; Department of Infectious Diseases, Clinic of Medicine, St. Olavs Hospital, 7006 Trondheim, Norway; Department of Chronic Disease Epidemiology, Yale University School of Public Health, New Haven, Connecticut, USA

## Abstract

**Importance:** Insomnia has been associated with reduced immune function and increased risk of infections and sepsis in observational studies. These studies are prone to bias, such as residual confounding. To further understand the causal relation between insomnia and sepsis risk we used a two-sample Mendelian randomization (MR) approach.

**Objective:** Is genetically predicted insomnia associated with risk of sepsis?

**Design:** Two-sample MR was performed to estimate the causal effect of genetically predicted insomnia on sepsis risk. Data was obtained from a genome-wide association study (GWAS) identifying 556 independent genetic variants (*R*^*2*^<0.01) strongly associated with insomnia (*P* < 5e-8). We conducted sensitivity analyses to address bias due to pleiotropy and sample overlap, along with mediation analyses.

**Setting:** Observational study using genetic variants as instrumental variables in large populations.

**Participants:** For insomnia, 2.4 million subjects of European ancestry from the UK Biobank and 23andMe. For sepsis, 462,918 subjects of European ancestry from the UK Biobank.

**Exposure:** Genetically predicted insomnia.

**Main Outcome and Measure:** Sepsis.

**Results:** A doubling in the population prevalence of genetically predicted insomnia was associated with an odds ratio of 1.42 (95% CI 1.23–1.63, *P* = 9.1e-7) for sepsis. Sensitivity analyses supported this observation. Three quarters of the effect was mediated through body mass index.

**Conclusions and Relevance:** The concordance between our findings and previous observational studies support of a causal role of genetically predicted insomnia in the risk of sepsis.

## Introduction

Insomnia is the most common sleep disorder^1^ and is associated with numerous adverse health outcomes.^2^ Current evidence suggests that insomnia can alter the immune function^3^ and possibly leave individuals suffering from insomnia more susceptible to infectious diseases. Sepsis is associated with high morbidity and mortality and is characterized by a dysregulated response to an infectious disease.^4^ It is therefore key to identify modifiable risk factors for sepsis that can be targets for preventive efforts. It has recently been reported that people with symptoms of insomnia have an elevated risk of bloodstream infection (BSI) – a condition closely linked to sepsis.^5^

Sleep disturbances could be influenced by factors or conditions associated with the risk of infectious diseases (e.g., diabetes, cardiovascular disease, adiposity and smoking),^2,5^ and the observed association between insomnia and infectious disease risk in previous studies may therefore be biased. Mendelian randomization (MR) is a powerful method using genetic variants as instruments for modifiable risk factors to reduce the influence of confounding and reverse causation.^6^ This approach mimics a randomized controlled trial by utilizing the principle of random allocation of genetic variants at meiosis and conception.

Our aim was to evaluate whether the previously reported association between insomnia and risk of systemic infectious diseases in observational studies also hold true when applying instrumental variable analyses using genetic instruments. We also aimed to estimate the proportion of the effect of genetically predicted insomnia on sepsis that is mediated through known risk factors of sepsis; i.e., body mass index (BMI), smoking and type 2 diabetes (T2DM).

## Method

We used a two-sample MR approach our objective. Single nucleotide polymorphisms (SNPs) were used as genetic instruments, and for each SNP we calculated the Wald ratio defined as the SNP-outcome association divided by the SNP-exposure association.^6^ For an instrument to be valid, it must be associated with the exposure, it cannot be associated with any confounder of the exposure-outcome association, and it cannot affect the outcome other than through the exposure.^6^

We obtained 556 genetic variants strongly associated (*P* <5e-8) with insomnia that were independent (*R*^2^<0.01) from a genome-wide association study (GWAS) evaluating 2.4 million subjects of European ancestry from the UK Biobank and 23andMe (Table 1).^7^ For the multivariable MR analyses (see below), we extracted genetic variants from relevant GWASs of BMI,^8^ T2DM^9^ and smoking status.^10^

**Table 1.**
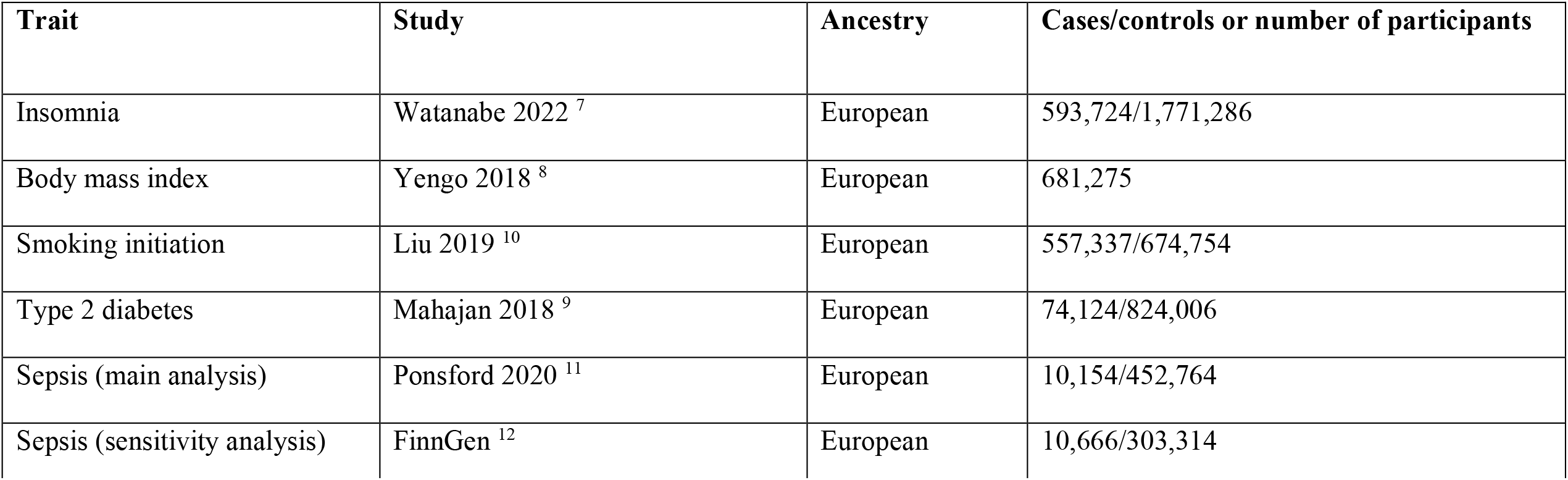
Overview of GWAS studies utilized in this study.

Genetic associations for sepsis were extracted from a GWAS in the UK Biobank^11^ including 10,154 sepsis cases and 452,764 controls, where cases were defined according to the explicit sepsis criteria defined in the most recent Global Burden of Disease study of sepsis.^4^ Because sample overlap between the exposure and outcome GWASs may bias the estimate towards the confounded estimate, we included a sensitivity analysis of sepsis (International Classification of Diseases, 10^th^ revision code A41) in FinnGen Release 8 including 10,666 cases and 303,314 controls.^12^

Prior to analyses, SNPs in the exposure and outcome GWASs were harmonized to evaluate the presence of the same allele. In the main analysis, using the inverse-variance weighted (IVW) method, we calculated the combined effect across the Wald ratios for all SNPs, putting more emphasis to the SNPs with the lowest variance. For the IVW estimate to be unbiased, all included instruments must be valid. Thus, we conducted sensitivity analyses using the weighted median, weighted mode and MR Egger regression that provide unbiased results even in the presence of some invalid instruments, but at the cost of lower statistical power.^6^

Lastly, we evaluated what proportion of the insomnia effect on sepsis that was mediated through three strong risk factors of sepsis; BMI, T2DM and smoking.^3,11,13^ Using the SNPs identified as genetic instruments for insomnia, we calculated the direct effect of insomnia on sepsis by conducting multivariable MR analyses with one of the three potential mediators at a time, and then all mediators combined. The univariable analyses yielded the total effect of genetically predicted insomnia. The proportion mediated was calculated as the direct effect divided by the total effect and subtracted from 1, and the standard errors were estimated using the propagation of error method.^6^

We used R (version 4.0.5) for data formatting and the TwoSampleMR package (version 0.5.6) for all analyses.^14^ All causal estimates were multiplied by 0.693 (= loge 2) to present the results as per doubling in the prevalence of insomnia. All data used were publicly available with relevant ethical approvals.

## Results

The genetic variants used in the main analysis explained 3.9% of the variance of insomnia. A genetically predicted doubling in the prevalence of insomnia was associated with an odds ratio (OR) of 1.42 (95% CI 1.23– 1.63, *P =* 9.1e-7) for sepsis (Figure 1). The weighted mode, weighted median and MR Egger analyses all supported the results of the main analysis. The result from the IVW analysis of risk of sepsis in FinnGen was similar to the analysis from UK Biobank (OR 1.25 [95% CI 1.09–1.44, *P* = 1.8e-3]). Finally, we found that most of the effect of genetically predicted insomnia on risk of sepsis was mediated through BMI, T2DM or smoking (Figure 2), but with a likely direct effect independent of these factors (OR 1.13 [95% CI 0.97–1.32, *P* = 3.4e-1]).

**Figure 1.**
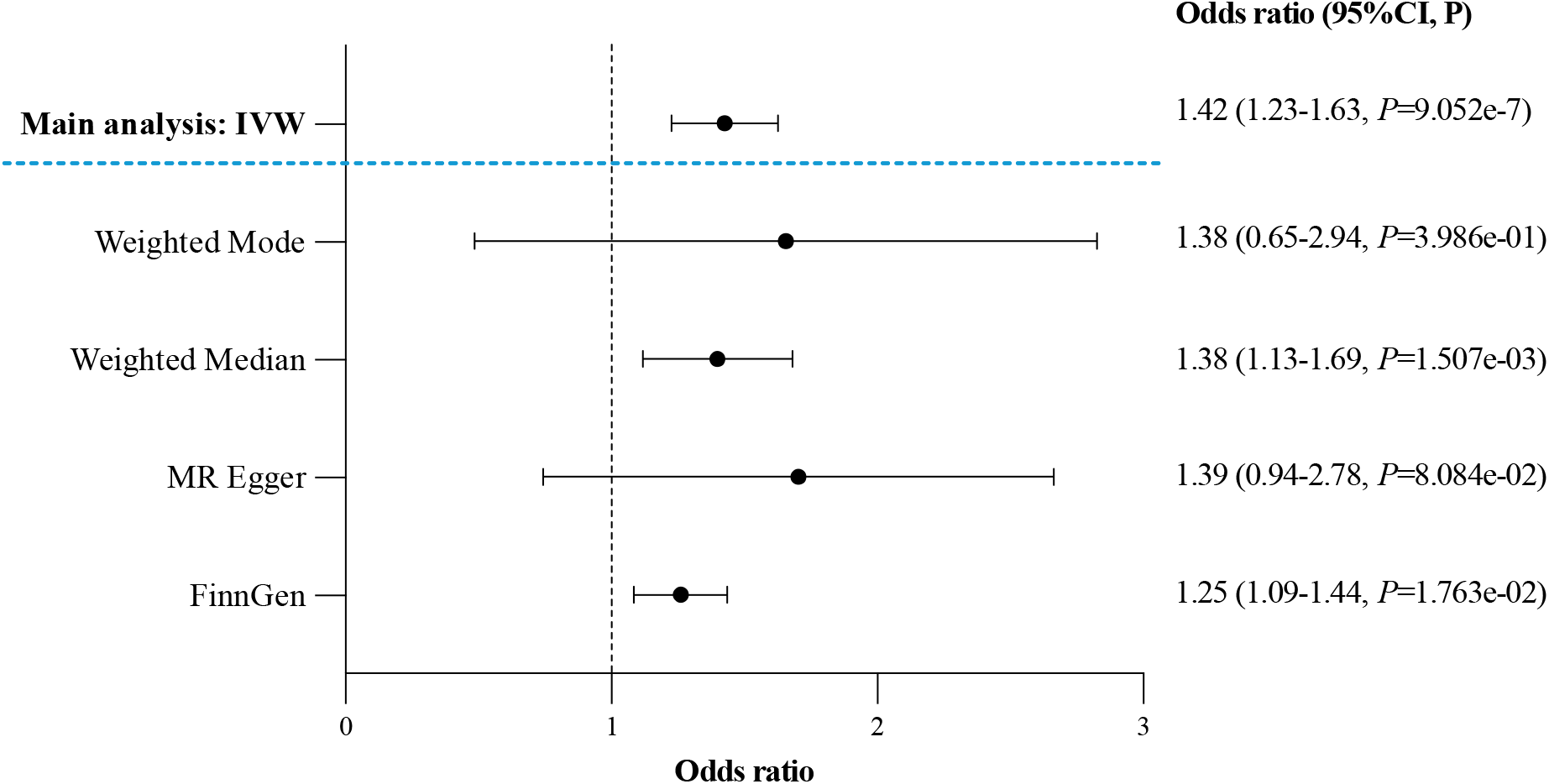
Mendelian randomization analyses of the association between genetically predicted insomnia and risk of sepsis. Odds ratios with 95% confidence intervals of risk of sepsis per genetically predicted doubling of the prevalence of insomnia. The FinnGen data was analyzed using IVW. Abbreviations; CI – confidence interval, IVW – inverse variance weighted, MR – Mendelian randomization.

**Figure 2.**
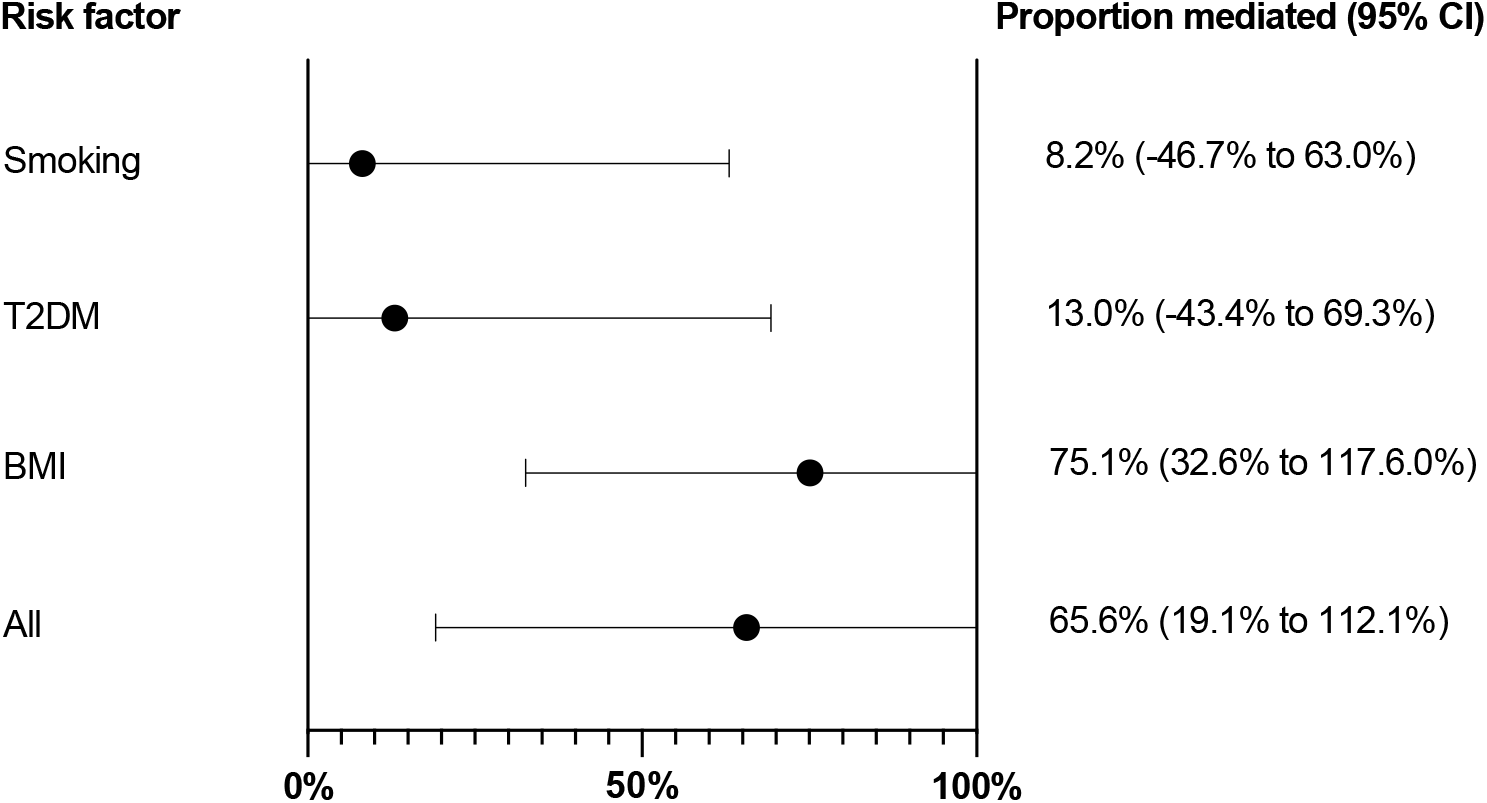
The proportion of the effect of genetically predicted insomnia on risk of sepsis that was mediated through smoking, type 2 diabetes, and body mass index. Abbreviations; BMI – Body mass index, CI – confidence interval, T2DM – Type 2 diabetes mellitus.

## Discussion

Our findings support a causal effect of genetically predicted insomnia on risk of sepsis. This is in line with previous research and results from observational data, reporting that insomnia increases the risk of altered immune response^3^ and bloodstream infection.^5^ We observed that large parts of the effect of genetically predicted insomnia on risk of sepsis were mediated through BMI, T2DM and smoking, which is in line with previous literature.^3,6,11,13^ Although the precise mechanisms of this association remain elusive, evidence suggests that sleep can promote the immune homeostasis through different mediators that both affect the innate and adaptive immune responses.^3^ Subpopulations of lymphocytes (e.g., total T-cells, CD4 T-cells and CD8 T-cells) was in a small study of 19 insomnia patients found to be fewer compared with healthy individuals.^15^ As summarized by Besedovsky et al., numerous studies have reported on insomnia altering immune function through various mechanisms (e.g., inflammatory cytokines, lymphocyte subsets and telomere length), suggesting insomnia in itself is linked to immune and inflammatory dysregulation.^3^

Our findings were strengthened by consistent findings across sensitivity analyses robust to pleiotropy, and when evaluating a separate outcome cohort with no overlap with the exposure GWAS. An important limitation of our study is that it only included individuals of European ancestry, and we encourage future studies to evaluate whether our findings replicate in other ancestry groups.

## Conclusion

The findings from this MR study are in accordance with those of previous observational studies and support a causal association between genetically predicted insomnia and risk of sepsis.

## Data Availability

All data produced are available online at

https://research.23andme.com/dataset-access/

https://www.ukbiobank.ac.uk/enable-your-research/apply-for-access

## Authors contribution

All authors have contributed significantly to the manuscript. All authors participated in designing the study. Marianne Thorkildsen wrote the manuscript. Tormod Rogne directed its implementation and performed the analyses. Jan Kristian Damås, Tom Ivar Lund Nilsen, Randi Marie Mohus and Lise Tuset Gustad contributed to the interpretation of the results and the final manuscript.

## Data access

All data is publicly available.

